# Predictors of COVID-19 vaccination uptake and reasons for decline of vaccination: a systematic review

**DOI:** 10.1101/2021.07.28.21261261

**Authors:** Petros Galanis, Irene Vraka, Olga Siskou, Olympia Konstantakopoulou, Aglaia Katsiroumpa, Daphne Kaitelidou

## Abstract

**Background:** Various COVID-19 vaccines with proven safety and effectiveness are available now but vaccine hesitancy remains a public threat. COVID-19 vaccines uptake appears to have an essential role in the successful control of the COVID-19 pandemic.

**Objective:** To examine predictors of COVID-19 vaccination uptake and reasons for decline of vaccination.

**Methods:** We followed the Preferred Reporting Items for Systematic Reviews and Meta-Analysis guidelines for this systematic review. We searched Medline, PubMed, Web of Science, Scopus, ProQuest, CINAHL, and a pre-print service (medRxiv) from inception to July 12, 2021. We used the following key-words: vaccin*, COVID-19, and uptake. We included all types of studies (quantitative, qualitative, and mixed methods) reporting COVID-19 vaccination uptake. The review protocol was registered with PROSPERO (CRD42021267460).

**Results:** Twelve studies met the inclusion and exclusion criteria. COVID-19 vaccination uptake ranged from 28.6% to 98% in the general population, while among healthcare workers ranged from 33.3% to 94.5%, and among patients ranged from 36% to 80%. The main predictors of COVID-19 vaccination uptake were male gender, white race, older age, higher socioeconomic status, higher self-perceived COVID-19 vulnerability, increased information about COVID-19 vaccines, and chronic illness. The most important reasons for decline of vaccination were concerns about the safety and effectiveness of vaccines, illness, medication, pregnancy, fertility, breastfeeding, religious reasons, ethical reasons, previous COVID-19 diagnosis, self-estimation that COVID-19 is not a severe disease, and limited knowledge about the vaccines.

**Conclusions:** Several factors affect COVID-19 vaccination uptake, while various reasons affect people’s decision to refuse to take a COVID-19 vaccine. These findings are essential to further enhance our understanding of COVID-19 vaccination uptake and design specific interventions. Given the high prevalence of COVID-19 vaccine hesitancy, our findings have major implications for the delivery of COVID-19 vaccination programmes in the public with special attention to people who are undecided or unlikely to take a COVID-19 vaccine.

## Background

From December 2020, several COVID-19 vaccines with proven efficacy and safety are being used worldwide (Baden et al., 2021; Logunov et al., 2021; Polack et al., 2020; Wu et al., 2021). Early real-world data have confirmed the effectiveness of COVID-19 vaccines against the severe acute respiratory syndrome coronavirus 2 (SARS-CoV-2) by reducing SARS-CoV-2 infection, COVID-19 cases, hospitalizations, unfavorable outcomes, and deaths among vaccinated individuals (Amit et al., 2021; Dagan et al., 2021; Haas et al., 2021; Hall et al., 2021; Vasileiou et al., 2021). Universal vaccination against SARS-CoV-2 is essential to safely achieve herd immunity and contain the COVID-19 pandemic (Khalife & VanGennep, 2021; Lacsa & Cordero, 2021; MacIntyre et al., 2021).

Willingness of the general population to accept COVID-19 vaccination is the first step to achieve a high rate of COVID-19 vaccination uptake and control the COVID-19 pandemic. However, vaccine hesitancy is one of the main obstacles to control the COVID-19 pandemic since some individuals refuse to take a COVID-19 vaccine (Jaca et al., 2021; Wiysonge et al., 2021). COVID-19 vaccination intention in the general population ranges from 27.7% to 97%, while lower rates are reported in Africa, Middle East, Russia, and several European countries (Al-Amer et al., 2021; Sallam, 2021). Similarly, a wide range of intention to accept COVID-19 vaccination (from 23.4% to 81.5%) is reported among healthcare workers with an overall proportion of 55.9% (Galanis et al., 2020; M. Li et al., 2021). Moreover, vaccine hesitancy is higher among healthcare workers and particularly among nurses than the general population due to concerns about the safety and effectiveness of vaccines (Al-Amer et al., 2021).

Several factors are shown to be associated with vaccine hesitancy, such as female gender, younger age, healthcare profession, low confidence in the government, concerns for safety, efficacy and effectiveness of COVID-19 vaccines, low levels of knowledge, negative information about COVID-19 vaccines, paranoid or conspiracy beliefs (Al-Amer et al., 2021; Butter et al., 2021; Galanis et al., 2020; M. Li et al., 2021; Murphy et al., 2021). On the other hand, morbidity, stronger vaccine confidence, seasonal influenza vaccination, positive attitude towards COVID-19 vaccines, fear against COVID-19, and high self-perceived risk of COVID-19 are positive predictive factors to accept a vaccine against COVID-19 (Galanis et al., 2020; M. Li et al., 2021).

Robust vaccination programs against SARS-COV-2 have been established throughout the world. Early studies have already investigated predictors of COVID-19 vaccination uptake but to date, no systematic review on this issue is published. Thus, the current systematic review aimed to examine predictors of COVID-19 vaccination uptake and reasons for decline of vaccination.

## Methods

### Data sources and strategy

A systematic review was conducted according to the Preferred Reporting Items for Systematic Reviews and Meta-Analysis (PRISMA) guidelines (Moher et al., 2009). We searched Medline, PubMed, Web of Science, Scopus, ProQuest, CINAHL, and a pre-print service (medRxiv) from inception to July 12, 2021. We used the following strategy in all fields: ((vaccin*) AND (COVID-19)) AND (uptake). The review protocol was registered with PROSPERO (registration number: CRD42021267460).

### Selection and eligibility criteria

After duplicates removal, we screened titles, abstracts, and full texts. Also, we examined reference lists of all relevant articles. Two independent authors performed study selection and a third, senior author resolved the differences. We included all types of studies (quantitative, qualitative, and mixed methods) reporting COVID-19 vaccination uptake. Also, we included studies that examine predictors of COVID-19 vaccination uptake and reasons for decline of vaccination. Studies published in English were eligible to be included. We excluded reviews, protocols, case reports, letters to the Editor, and editorials.

### Data extraction and quality assessment

Two reviewers independently extracted the following data from the studies: authors, country, data collection time, sample size, age, population, study design, sampling method, response rate, percentage of COVID-19 vaccination uptake, predictors of COVID-19 vaccination uptake, reasons for decline of COVID-19 vaccination, and type of publication (journal or pre-print service). We appraised each study’s quality using the Joanna Briggs Institute critical appraisal tool for cross-sectional studies (Santos et al., 2018).

## Results

### Identification and selection of studies

As shown in Figure 1, our initial search yielded 1682 records after duplicates removal. After following the inclusion and exclusion criteria, 12 articles were identified.

**Figure 1.**
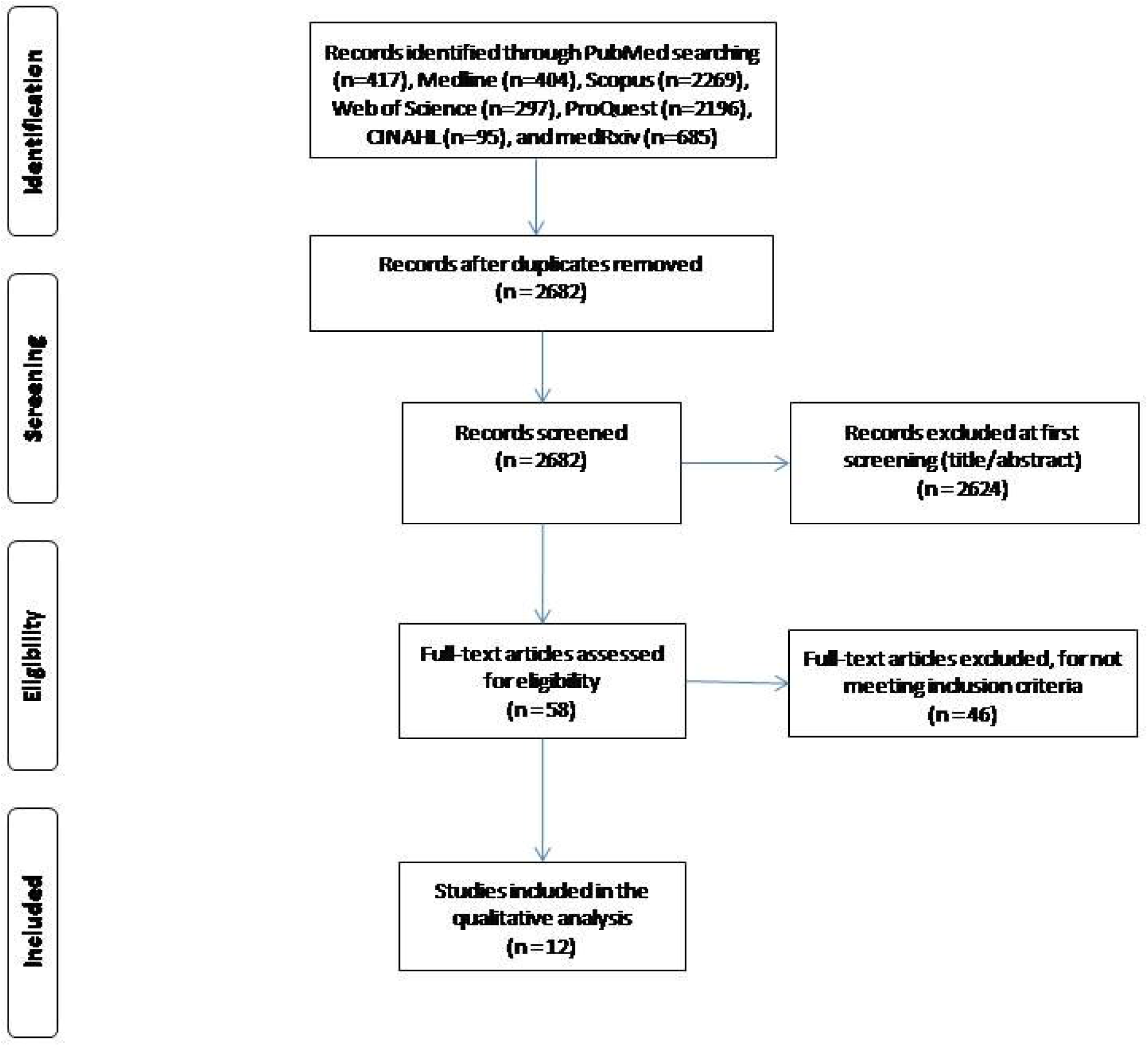
Flowchart of the literature search according to the Preferred Reporting Items for Systematic Reviews and Meta-Analysis.

### Characteristics of the studies

Main characteristics of the 12 studies included in this systematic review are presented in Table 1. Four studies were conducted in the USA (Gharpure et al., 2021; McCabe et al., 2021; Pacella-LaBarbara et al., 2021; Schrading et al., 2021), four studies in United Kingdom (Gibbon et al., 2021; Glampson et al., 2021; Martin et al., 2021; The OpenSAFELY Collaborative et al., 2021), one study in China (Xu et al., 2021), one study in Kingdom of Saudi Arabia (Barry et al., 2021), and one study in Poland (Malesza & Bozym, 2021). Also, one study included participants from the USA and United Kingdom (L. Nguyen et al., 2021). Data collection time among studies ranged from December 2020 (Barry et al., 2021; Gharpure et al., 2021) to May 2021 (McCabe et al., 2021), while the response rate ranged from 78.3% (Malesza & Bozym, 2021) to 100% (Gibbon et al., 2021). All studies were cross-sectional and a convenience sampling method was used. Sample size ranged from 85 (Gibbon et al., 2021) to 20,852,692 participants (The OpenSAFELY Collaborative et al., 2021). Five studies were published in journals (Gharpure et al., 2021; Gibbon et al., 2021; Pacella-LaBarbara et al., 2021; Schrading et al., 2021; Xu et al., 2021) and seven studies in pre-print services (Barry et al., 2021; Glampson et al., 2021; Malesza & Bozym, 2021; Martin et al., 2021; McCabe et al., 2021; L. Nguyen et al., 2021; The OpenSAFELY Collaborative et al., 2021).

**Table 1.**
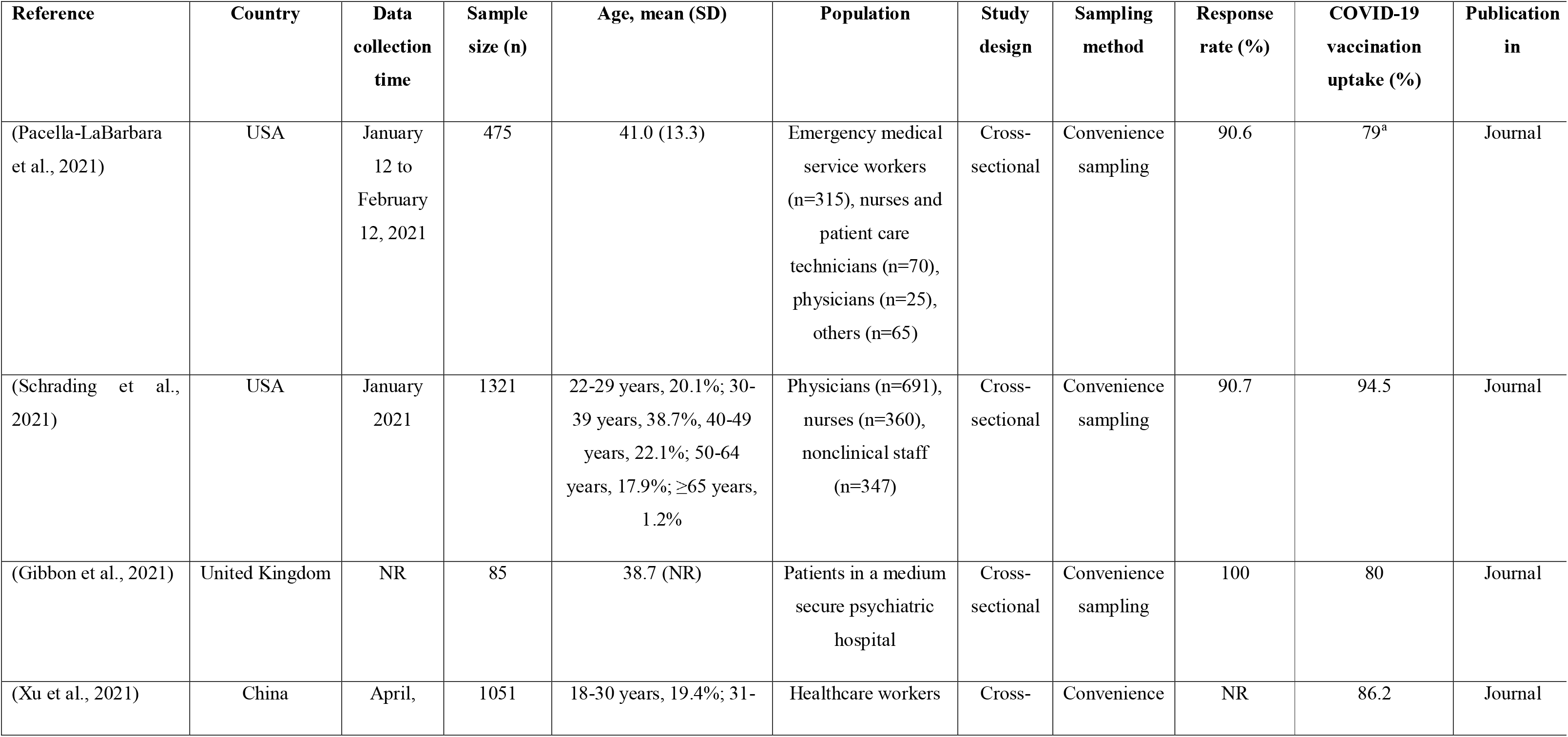
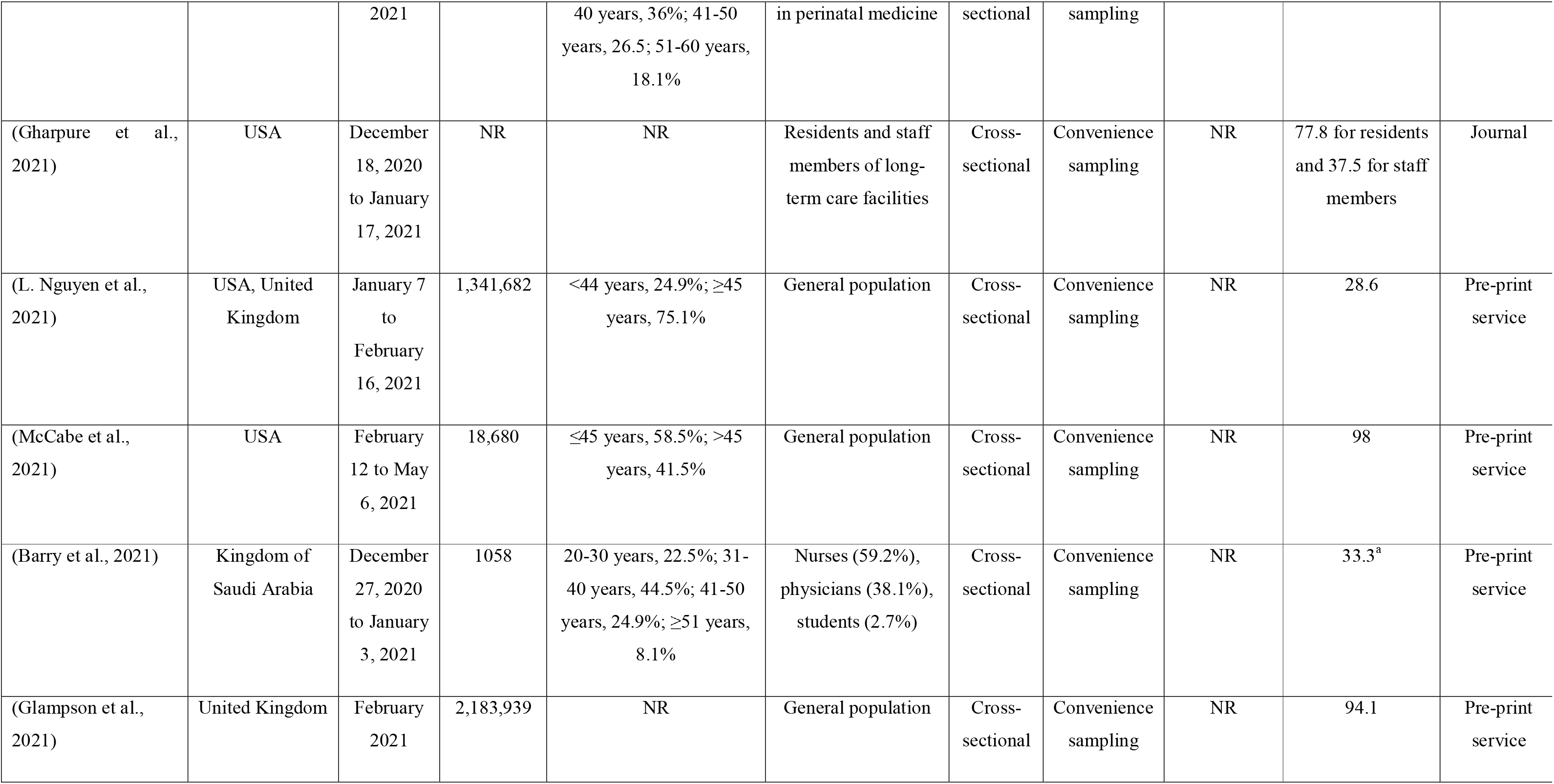
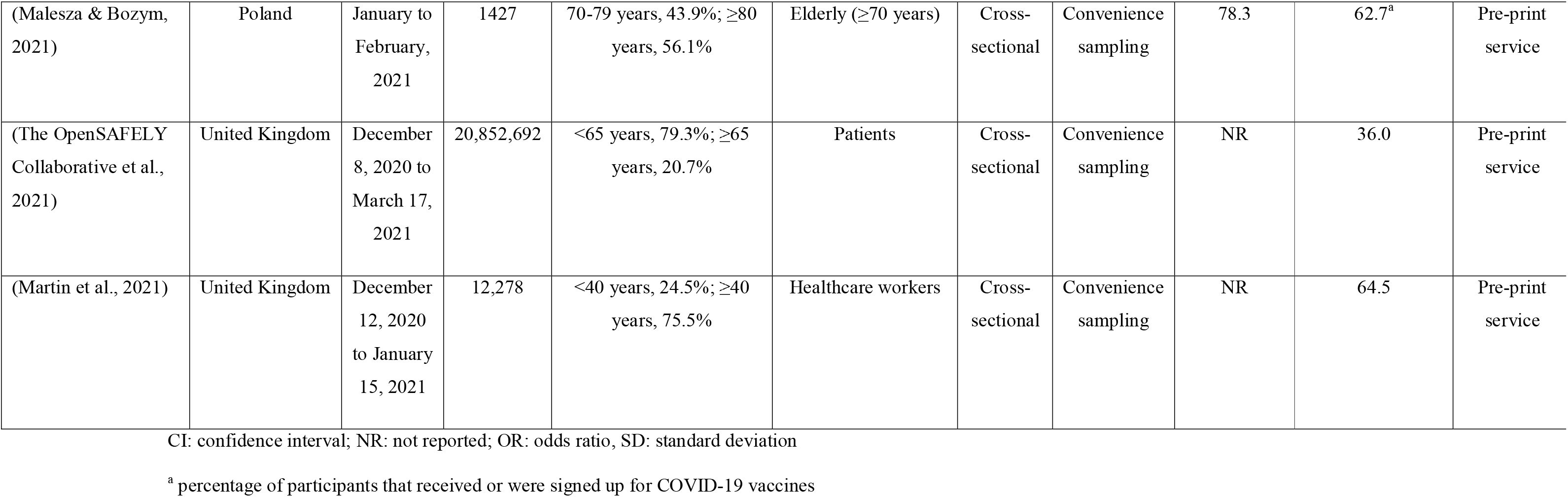
Overview of the studies included in this systematic review.

Study population included general population (Glampson et al., 2021; McCabe et al., 2021; L. Nguyen et al., 2021), healthcare workers (Barry et al., 2021; Martin et al., 2021; Pacella-LaBarbara et al., 2021; Schrading et al., 2021; Xu et al., 2021), patients (Gibbon et al., 2021; The OpenSAFELY Collaborative et al., 2021), elderly (Malesza & Bozym, 2021), and residents and staff members of long-term care facilities (Gharpure et al., 2021). COVID-19 vaccination uptake ranged from 28.6% (L. Nguyen et al., 2021) to 98% in the general population (McCabe et al., 2021), among healthcare workers ranged from 33.3% (Barry et al., 2021) to 94.5% (Schrading et al., 2021), and among patients ranged from 36% (The OpenSAFELY Collaborative et al., 2021) to 80% (Gibbon et al., 2021). Also, COVID-19 vaccination uptake was 62.7% in a sample of elderly (Malesza & Bozym, 2021), and 77.8% among residents of long-term care facilities (Gharpure et al., 2021).

Eight studies did not report data regarding response rate (Barry et al., 2021; Gharpure et al., 2021; Glampson et al., 2021; Martin et al., 2021; McCabe et al., 2021; L. Nguyen et al., 2021; The OpenSAFELY Collaborative et al., 2021; Xu et al., 2021), two regarding age (Gharpure et al., 2021; Glampson et al., 2021), one regarding data collection time (Gibbon et al., 2021), and one regarding sample size (Gharpure et al., 2021).

### Quality assessment

Quality assessment of cross-sectional studies included in this review is shown in Table 3. Quality was good in seven studies (Barry et al., 2021; Malesza & Bozym, 2021; Martin et al., 2021; McCabe et al., 2021; L. Nguyen et al., 2021; Pacella-LaBarbara et al., 2021; Xu et al., 2021), moderate in four studies (Gharpure et al., 2021; Glampson et al., 2021; Schrading et al., 2021; The OpenSAFELY Collaborative et al., 2021), and poor in one study (Gibbon et al., 2021). Five studies did not identify confounding factors (Gharpure et al., 2021; Gibbon et al., 2021; Glampson et al., 2021; Schrading et al., 2021; The OpenSAFELY Collaborative et al., 2021), three studies did not describe in detail the study subjects and the setting (Gharpure et al., 2021; Gibbon et al., 2021; Glampson et al., 2021), two studies did not measure the exposure in a valid and reliable way (Gharpure et al., 2021; Gibbon et al., 2021), and two studies did not use the appropriate statistical analysis (Gibbon et al., 2021; Schrading et al., 2021) (Table 2).

**Table 2.** Quality of cross-sectional studies included in this systematic review.

|  | (Pacella-LaBarbara et al., 2021) | (Schrading et al., 2021) | (Gibbon et al., 2021) | (Xu et al., 2021) | (Gharpure et al., 2021) | (L. Nguyen et al., 2021) | (McCabe et al., 2021) | (Barry et al., 2021) | (Glampson et al., 2021) | (Malesza & Bozym, 2021) | (The OpenSAFELY Collaborative et al., 2021) | (Martin et al., 2021) |
| --- | --- | --- | --- | --- | --- | --- | --- | --- | --- | --- | --- | --- |
| 1. Were the criteria for inclusion in the sample clearly defined? | √ | √ | √ | √ | √ | √ | √ |  | √ | √ | √ | √ |
| 2. Were the study subjects and the setting described in detail? | √ | √ |  | √ |  | √ | √ | √ |  | √ | √ | √ |
| 3. Was the exposure measured in a valid and reliable way? | √ | √ |  | √ |  | √ | √ | √ | √ | √ | √ | √ |
| 4. Were objective, standard criteria used for measurement of the condition? | √ | √ | √ | √ | √ | √ | √ | √ | √ | √ | √ | √ |
| 5. Were confounding factors identified? | √ |  |  | √ |  | √ | √ | √ |  | √ |  | √ |
| 6. Were strategies to deal with confounding factors stated? | √ |  |  | √ |  | √ | √ | √ |  | √ |  | √ |
| 7. Were the outcomes measured in a valid and reliable way? | √ | √ | √ | √ | √ | √ | √ | √ | √ | √ | √ | √ |
| 8. Was appropriate statistical analysis used? | √ |  |  | √ | √ | √ | √ | √ | √ | √ | √ | √ |
| **Total quality** | Good | Moderate | Poor | Good | Moderate | Good | Good | Good | Moderate | Good | Moderate | Good |

**Table 3.** Predictors of COVID-19 vaccination uptake and reasons for decline of vaccination.

| Reference | Predictors of COVID-19 vaccination uptake | Reasons for decline of COVID-19 vaccination |
| --- | --- | --- |
| (Pacella-LaBarbara et al., 2021) | Vaccination uptake was higher among: (a) males (OR=2.94; 95% CI=1.1 to 4.17; p-value=0.02), (b) participants without a history of COVID-19 infection (OR=1.82; 95% CI=1.02 to 3.23; p-value=0.041), (c) participants with advanced degree (OR=3.53; 95% CI=1.16 to 10.77; p-value=0.026), and (d) participants with higher perceived COVID-19 vulnerability (OR=1.99; 95% CI=1.37 to 2.90; p-value<0.001). |  |
| (Schrading et al., 2021) | Vaccination uptake was higher: (a) among physicians (94.5%) than nurses (77.7%) and nonclinical staff (76.5%), (b) among males (93.5%) than females (81.6%), and (c) among white individuals (88.5%) than black individuals (65.4%). | Safety of vaccines (45.4%), health condition (13.5%), previous COVID-19 diagnosis (13.5%), pregnancy/fertility (11.9%), religious/ethical/personal reasons (8.6%), concerns about the vaccines efficacy (8.1%), and logistics/scheduling (4.3%). |
| (Gibbon et al., 2021) | Vaccination uptake was higher among white British patients (83.1%) than Black Asian minority ethnic patients (70%). | Concerns about the safety of vaccines (29.4%) and patients' perception of having a low risk from the COVID-19 (23.5%). |
| (Xu et al., 2021) |  | Concerns about the safety of vaccines, pregnancy, and breastfeeding |
| (L. Nguyen et al., 2021) | Vaccination uptake was higher among white participants than Black participants (OR=1.41; 95% CI=1.27 to 1.56; p-value<0.001). | Concerns about the safety of vaccines, limited knowledge about the vaccines, religious/ethical/personal reasons, illness/medication, pregnancy, and breastfeeding. |
| (McCabe et al., 2021) | Vaccination uptake was lower among black participants, younger participants, healthcare workers, participants from lower income households, and participants of areas with lower population density (p<0.05 in all cases). |  |
| (Barry et al., 2021) | Vaccination uptake was higher among: (a) males (OR=3.48; 95% CI=2.49 to 4.85; p-value<0.001), (b) participants with age >40 years (OR=1.02; 95% CI=1.002 to 1.04; p-value=0.032), (c) native-born participants (OR=1.92; 95% CI=1.36 to 2.69; p-value<0.001), (d) healthcare workers in intensive care unit (OR=1.49; 95% CI=1.08 to 2.06; p-value=0.041), (e) healthcare workers in university hospitals (OR=1.87; 95% CI=1.38 to 2.53; p-value<0.001), and (f) participants that did not use social media as a source of information (OR=4.83; 95% CI=2.82 to 7.57; p-value=0.001). |  |
| (Glampson et al., 2021) | Vaccination uptake was lower among black individuals. Negative association between deprivation and vaccination uptake (correlation coefficient=-0.94, p-value<0.01). |  |
| (Malesza & Bozym, 2021) | Vaccination uptake was higher among: (a) participants living with others (OR=3.13; 95% CI=2.03 to 4.26; p-value<0.05), (b) participants having a high rank occupation (OR=1.79; 95% CI=1.33 to 2.15; p-value<0.05), (c) participants being able to access medical services by driving or walking (OR=1.92; 95% CI=1.45 to 2.76; p-value<0.05), (d) participants suffering from a chronic illness (OR=2.98; 95% CI=2.05 to 4.01; p-value<0.05), (e) participants that physicians explained the importance of COVID-19 vaccination (OR=4.23; 95% CI=2.90 to 5.75; p-value<0.05), and (f) participants that physicians explained the possible side effects of COVID-19 vaccination (OR=3.48; 95% CI=2.03 to 4.85; p-value<0.05). | Concerns about the safety of vaccines (91.4%), self-estimation that COVID-19 is not a severe disease (75.4%), concerns about the effectiveness of COVID-19 vaccination (66.7%), bad experiences of vaccines among family members/friends (57.1%), medical reasons (53%), inconvenience (30.8%), and personal bad experiences of vaccines (19.2%). |
| (Martin et al., 2021) | Vaccination uptake was higher among: (a) males (p-value<0.001), (b) older participants (p-value<0.001), (c) white participants (p-value<0.001), (d) allied health professionals and administrative/executive staff (p-value<0.001), and (e) participants without a history of COVID-19 infection (p-value<0.001). |  |

### Predictors of COVID-19 vaccination uptake and reasons for decline of vaccination

Nine studies investigated predictors of COVID-19 vaccination uptake (Barry et al., 2021; Gibbon et al., 2021; Glampson et al., 2021; Malesza & Bozym, 2021; Martin et al., 2021; McCabe et al., 2021; L. Nguyen et al., 2021; Pacella-LaBarbara et al., 2021; Schrading et al., 2021), while five studies searched for reasons for decline of COVID-19 vaccination (Gibbon et al., 2021; Malesza & Bozym, 2021; L. Nguyen et al., 2021; Schrading et al., 2021; Xu et al., 2021) (Table 3).

We found that several factors affect COVID-19 vaccination uptake. In particular, seven studies found that white individuals had the highest rate of vaccination uptake, while black individuals have the lowest rate (Gibbon et al., 2021; Glampson et al., 2021; Martin et al., 2021; McCabe et al., 2021; L. Nguyen et al., 2021; Schrading et al., 2021; Xu et al., 2021). Also, Barry et al. (2021) found that native-born participants were vaccinated for COVID-19 more often than immigrants. Gender and age were predictors of COVID-19 vaccination uptake since four studies (Barry et al., 2021; Martin et al., 2021; Pacella-LaBarbara et al., 2021; Schrading et al., 2021) found that uptake was higher among males and three studies (Barry et al., 2021; Martin et al., 2021; McCabe et al., 2021) found that uptake was higher among older participants. Higher education level (Schrading et al., 2021), higher income (McCabe et al., 2021), and higher rank occupation (Malesza & Bozym, 2021) were related with higher levels of COVID-19 vaccination uptake. COVID-19 vaccination uptake was more likely in physicians (Schrading et al., 2021), in allied health professionals and administrative/executive staff (Martin et al., 2021), and in healthcare workers in university hospitals and intensive care units (Barry et al., 2021). According to McBabe et al. (2021), general population was vaccinated more often than healthcare workers. Participants without a history of COVID-19 infection (Martin et al., 2021; Pacella-LaBarbara et al., 2021), participants with higher self-perceived COVID-19 vulnerability (Pacella-LaBarbara et al., 2021), and participants that were more informed about COVID-19 vaccines (Malesza & Bozym, 2021) were more likely to be vaccinated. Malesza & Bozym (2021) found that vaccination uptake was higher among participants living with others and those with a chronic illness.

The most important reasons for decline of vaccination were concerns about the safety and effectiveness of vaccines, illness, medication, pregnancy, fertility, breastfeeding, religious reasons, ethical reasons, previous COVID-19 diagnosis, self-estimation that COVID-19 is not a severe disease, bad experiences of vaccines among family members/friends, and limited knowledge about the vaccines (Gibbon et al., 2021; Malesza & Bozym, 2021; L. Nguyen et al., 2021; Schrading et al., 2021; Xu et al., 2021).

## Discussion

To our knowledge, this is the first systematic review that examines predictors of COVID-19 vaccination uptake and reasons for decline of vaccination. Twelve papers met our inclusion and exclusion criteria and COVID-19 vaccination uptake ranged from 28.6% to 98% in the general population, while among healthcare workers ranged from 33.3% to 94.5%.

According to our systematic review, COVID-19 vaccination uptake was higher among white individuals than black individuals. This result echoes the findings of research which shows a higher rate of COVID-19 vaccine hesitancy in ethnic minorities, particularly in Black and Asian groups (Funk & Tyson, 2021; Hamel et al., 2021; K. H. Nguyen et al., 2021; Ruiz & Bell, 2021). Lack of trust in the governments, lower self-perception of the risk of COVID-19, concerns about the safety of vaccines, religious factors, fear of adverse reactions, and lower socioeconomic status considered to be barriers to vaccine uptake in ethnic minorities (Forster et al., 2017; Gamble, 1997; Mills et al., 2020). Moreover, health-protective behaviors during the COVID-19 pandemic such as mask usage is shaped by an insensitivity to deaths among Black and Latin Americans (Franz et al., 2021). In particular, mask wearing increased when death rates among White Americans relative to death rates among Black and Latin Americans increased. Ethnic minority groups are high-risk groups for infection with SARS-CoV-2 and adverse outcomes from COVID-19 and further consideration should be given to how the COVID-19 vaccination uptake can be improved in people from ethnic minorities (Martin, Jenkins, et al., 2020; Martin, Patel, et al., 2020; Sze et al., 2020; Voysey et al., 2021).

We found that male gender and older age were related with increased COVID-19 vaccination uptake. This finding is plausible since it is well known that older age is a significant predictor of COVID-19 mortality (Mehraeen et al., 2020; Sepandi et al., 2020; Yanez et al., 2020). Also, males are generally less likely to report COVID-19 vaccine hesitancy and more likely to accept a COVID-19 vaccine than females (Dror et al., 2020; Gagneux-Brunon et al., 2021; Khubchandani et al., 2021; Lazarus et al., 2021; Malik et al., 2020). Additionally, male patients require more often intensive care unit admission and show higher mortality compare to females (Bienvenu et al., 2020; Peckham et al., 2020). Probably, older males confront COVID with more anxiety, distress and fear resulting on a higher vaccination uptake. Moreover, females’ COVID-19 vaccine hesitancy may also be related with limited knowledge regarding issues such as pregnancy, fertility and breastfeeding (L. Nguyen et al., 2021; Schrading et al., 2021; Xu et al., 2021).

Our review identified that participants with higher self-perceived COVID-19 vulnerability were more likely to be vaccinated. This finding confirms that risk perception with regards to COVID-19 is critical to vaccination behavior (Caserotti et al., 2021). In particular, as risk perception and worry about contracting COVID-19 increase, so does the intention to accept a COVID-19 vaccine (Caserotti et al., 2021; Glöckner et al., 2020; Ward et al., 2020). Individuals that consider COVID-19 as a severe disease are more intent on taking a COVID-19 vaccine (Karlsson et al., 2021). Similarly, according to our review, self-estimation that COVID-19 is not a severe disease was a reason for people to refuse to have a COVID-19 vaccine (Malesza & Bozym, 2021). Interestingly, people were more likely to accept a COVID-19 vaccine during the lockdown periods when they felt more vulnerable (Brooks et al., 2020; Z. Li et al., 2020; Moccia et al., 2020). Moreover, the low self-perceived risk of contracting COVID-19 is related with low vaccination rate (Karlsson et al., 2021). This finding is confirmed by our review since we found that participants without a history of COVID-19 infection were more likely to be vaccinated, potentially due to a high self-perceived risk of contracting SARS-CoV-2 and/of negative clinical outcomes. Also, we found that previous COVID-19 diagnosis was a reason for decline of vaccination. Probably, past COVID-19 patients feel protected against the disease and perceive a low risk of contracting SARS-CoV-2 again (Pacella-LaBarbara et al., 2021).

Also, we found that participants with a chronic illness were vaccinated more often than healthy participants. Worry about contracting COVID-19 increases the intention to accept a COVID-19 vaccine and probably people with medical conditions feel more fear, stress, worry, psychological distress, and anxiety during the COVID-19 pandemic (Blix et al., 2021; Salari et al., 2020; Xiong et al., 2020). Illness and medication cause a negative attitude towards COVID-19 vaccination since patients have concerns about the safety of vaccines and consider their illness as a contradiction of vaccination (L. Nguyen et al., 2021). In general, concerns about the safety and effectiveness of COVID-19 vaccines is the most important reason for decline of vaccination (Gibbon et al., 2021; Malesza & Bozym, 2021; L. Nguyen et al., 2021; Schrading et al., 2021; Wu et al., 2021). Similarly, according to our review, females fear that COVID-19 vaccines may cause problems in pregnancy, fertility, and breastfeeding and then refuse to take a COVID-19 vaccine (L. Nguyen et al., 2021; Schrading et al., 2021; Xu et al., 2021).

Moreover, concerns about safety, effectiveness, side-effects, and efficacy of COVID-19 vaccines and general distrust are related with hesitancy in COVID-19 vaccines uptake in the community (Freeman et al., 2020). According to WHO, vaccine hesitancy is a top ten global health threat in 2019 since it is one of main obstacles to control vaccine preventable diseases such as COVID-19 (Jaca et al., 2021; Wiysonge et al., 2021; World Health Organization, 2020). Moreover, vaccine hesitancy among healthcare workers warrants particular attention since they could put patients at risk and also their negative attitude towards COVID-19 vaccination may significantly decrease public uptake of COVID-19 vaccines (Gadoth et al., 2021; Schaffer DeRoo et al., 2020). This finding was confirmed by this review since we found that general population was vaccinated more often than healthcare workers and hesitant workers were more likely to change their opinion regarding COVID-19 vaccination than healthcare workers (McCabe et al., 2021). Healthcare workers are the most important predictor of vaccine acceptance in the general population and a strong recommendation from them could improve significantly vaccine acceptance (Allison et al., 2013; Dorell et al., 2011; Opel et al., 2013).

As we found in our review, limited knowledge about the vaccines decreases the probability to take a COVID-19 vaccine (L. Nguyen et al., 2021). Various cultural, social, political, personal, and religious factors contribute to vaccine hesitancy creating a complex issue (Kestenbaum & Feemster, 2015). In that case, knowledge is essential to decrease vaccine hesitancy since it is well known that misinformation and negative stories about vaccine safety in the social media and news create mistrust in biomedical sciences and negative attitudes towards vaccination (Dubé et al., 2013; Gust et al., 2005). Moreover, higher socioeconomic status was related with higher probability of COVID-19 vaccination uptake (Malesza & Bozym, 2021; McCabe et al., 2021; Schrading et al., 2021). Probably, higher education level and income are related with higher level of knowledge regarding vaccines, more trust in biomedical research and governments, and higher probability to afford the logistics regarding a vaccine uptake. Education programs have already proved effective to increase influenza vaccination rate and may be used as a guide in case of COVID-19 vaccination also (Black et al., 2018).

### Limitations

Our systematic review is subject to several limitations. Firstly, five out of 12 studies was of poor or moderate quality, while more than the half of studies was published in pre-print services without a peer-review process. Secondly, studies were conducted mainly in the USA and United Kingdom and thus the results could not be generalized. Thirdly, data collection time among studies ranged from December 2020 to May 2021. Availability of COVID-19 vaccines and knowledge regarding these vaccines are increasing significantly on an ongoing basis and people’s attitudes towards COVID-19 vaccination could be changed. Moreover, all studies in our review were cross-sectional and thus causal inferences are impossible. Also, only nine studies investigated predictors of COVID-19 vaccination uptake and five studies investigated reasons for decline of COVID-19 vaccination. Additional research is needed to understand as soon as possible the factors that influence people’s decision to take a COVID-19 vaccine. For instance, no study until now has investigated psychological factors that could affect people’s attitudes towards COVID-19 vaccination uptake. Moreover, we should examine whether additional sociodemographic, infection-related and social media variables are related with COVID-19 vaccination uptake.

### Conclusions

Several factors affect COVID-19 vaccination uptake, while there are various reasons for decline of vaccination. For instance, males, older and white people take more often a COVID-19 vaccine. These findings are essential to further enhance our understanding of COVID-19 vaccination uptake and design specific interventions. Information campaigns with regards to COVID-19 vaccination are of paramount importance and should promote group strategies, focusing on informing the public about the safety of COVID-19 vaccines and addressing the concerns of people who are undecided or unlikely to take a COVID-19 vaccine. Also, these campaigns should offer reassurance, especially to groups that have more concerns about the safety of COVID-19 vaccines, e.g. females, young adults, people from ethnic minorities, people with limited knowledge about the vaccines etc. Large proportions of these populations are undecided and reliable COVID-19 vaccination information should be provided tailored to the needs of each sub-group. Given the high prevalence of COVID-19 vaccine hesitancy, our findings have major implications for the delivery of COVID-19 vaccination programmes in the public with special attention to the groups identified in this review.

## Data Availability

Data will be available after reasonable request

## Notes

### Competing Interest Statement

The authors have declared no competing interest.

### Funding Statement

none to declare

### Author Declarations

We performed a systematic review. Thus there is no need for approval from an ethics committee.

